# Improving Adherence to Pulmonary Embolism Evaluation Consensus Guidelines: A Multimodal Approach

**DOI:** 10.1101/2024.02.08.24302549

**Authors:** E Ethan D’Silva, Syed Zaidi, Phil Ramis, Eric M. Rohren

## Abstract

**Objective:** Despite the publication of multiple widely accepted guidelines, Computed Tomography pulmonary angiography (CTPA) remains overutilized in the emergency (ED) setting for evaluating suspected pulmonary embolism (PE). We developed and evaluated a multimodal program that aimed to improve ordering provider adherence to CTPA use guidelines, and consequently positivity rate.

**Methods:** We retrospectively identified all CTPA studies ordered for suspected PE over three years for each provider at a single-center ED, and reviewed radiology reports to determine PE positivity. We then implemented a program that included an education session, individualized performance feedback, and discussion of possible incentives to improve guideline adherence. We prospectively collected CTPA results for the following year. Our primary outcome was the difference in positivity rate between the final preintervention year and the post-intervention year. Our secondary outcome was individual provider positivity rate between the pre-intervention and post-intervention period. We compared the primary outcome using chi-squared testing, defining statistical significance as P<0.05.

**Results:** We identified 4265 CTPA studies performed during the pre-intervention period, ordered by 25 providers. 1582 studies were ordered during the final year, with an 11.00% positivity rate (174/1582). In the post-intervention year, 1339 studies were ordered, with a significantly greater positivity rate of 13.67% (183/1339, P = 0.028). For our secondary outcome, mean provider positivity rate was 11.49% during the preintervention period, and increased to 14.24% in the post-intervention period.

**Discussion:** We successfully implemented a multimodal program for ED providers that significantly increased the positivity rate of CTPAs ordered for suspected PE evaluation.

## Introduction

Computed Tomography pulmonary angiography (CTPA) is the gold standard for evaluating suspected pulmonary embolism (PE), yet despite receiving considerable, continued attention during the past two decades, over-testing in the United States remains remarkably prevalent^1,2^. In one decade alone, CTPA use for suspected PE has increased between 3 and 13 times, and a 2017 analysis has shown that average CTPA utilization to diagnose PE is over five times greater in the US compared to the rest of the world^3^. While clinically valuable, CTPA studies are costly and come with significant medical risks, including radiation and contrast exposure. Moreover, CT examinations often reveal incidental findings of doubtful clinical significance, which may lead to additional workup, compounding both patient anxiety and financial burden.

To reduce overutilization, several clinical prediction rules (CPRs) have been developed to encourage more appropriate CTPA ordering, including the Wells and Geneva scores. Use of these guidelines in the emergency settings has been endorsed by multiple major physician groups, including the American College of Chest Physicians, the American Thoracic Society, the American College of Emergency Physicians, and Choosing Wisely^3^. Yet, guideline adherence during the past decade remains limited, with a substantial portion of CTPAs for suspected PE obtained outside of recommendation^4^.

In this study, we developed a multimodal program with a low resource footprint to improve appropriate use of CTPA when diagnosing suspected PE in the emergency setting. The program consisted of preparing individualized CTPA use analytics to providers, as well as a high-value care educational intervention. We implemented and evaluated this program with ordering clinicians at a single-institution emergency department. We hypothesized that such an intervention would improve CTPA positivity rate, by increasing adherence to currently accepted guidelines.

## Methods

### Data Collection

We identified all CTPA studies ordered by emergency department physicians credentialed at the participating hospital from January 1, 2012 to December 31, 2014: pre-intervention baseline years 1 - 3. We then further stratified these orders by provider. Using mPower (Nuance, Burlington, Massachusetts), a commercially available, HIPAA-compliant natural language processing software, we reviewed radiologist free-text reports to determine PE positivity for each study. We defined PE positive as presence of either isolated subsegmental PE or multiple PE without right heart strain. Using these criteria we calculated an overall PE positivity rate for each baseline year, as well as for each emergency department provider during this baseline period. This study was approved by the institutional review board of Aultman Hospital. All patient-specific data was de-identified during analysis. Given the de-identified nature of the study, waiver of consent was obtained.

### Intervention

Once baseline was established, a didactic, interactive education session was implemented for ordering providers. The session was conducted by emergency medicine and radiology physicians at the participating hospital. Session content consisted of the following three components.

First, session educators reviewed accepted clinical prediction rules used to determine pre-test PE probability: Gestalt criteria, Wells criteria, and Geneva criteria. Educators additionally reviewed the pulmonary embolism rule out criteria (PERC). During this review, ordering providers were able to ask questions to clarify their understanding. All ordering provider questions regarding the reviewed clinical tools were answered.

Second, each ordering provider was given a report of their individual CTPA use, including quantity of orders placed during the baseline period, and cumulative positivity rate. They were further provided with a report of aggregate CTPA use by the entire department during the baseline period, along with positivity rate. Third, educators and participants discussed additional incentives, such as financial bonuses, that could be implemented in the emergency department to improve adherence to the clinically accepted guidelines. Notably however, no incentives were implemented during the postintervention period.

### Evaluation and Outcomes

Following the educational intervention, we prospectively analyzed CTPA ordering practices at the department during a one-year post-intervention period: January 1, 2015 to December 1, 2015, in the same manner as previously described. No potentially confounding institutional changes were implemented in the post-intervention period. We then compared the results from the post-intervention period to those from the baseline period. Our primary outcome was the difference in positivity rate between the final preintervention year and the post-intervention year. As a secondary outcome, we compared the center and spread of individual provider positivity rates during the baseline period to those of the post-intervention period.

### Statistical Analysis

Statistical analysis for this study was performed using Microsoft Excel (Microsoft, Redmond, Washington) and R version 4.2.0 (R Foundation for Statistical Computing, Vienna, Austria). We then used χ^2^ or Fisher’s exact test, as appropriate, to compare our primary outcome, defining statistical significance as P < 0.05. For our secondary outcomes, we assessed normality using Shapiro-Wilkes testing. If the data were normally distributed, we compared mean and standard deviation. If the data were not normally distributed, median and IQR were used.

## Results

### Study Cohort and Primary Outcome

We identified 4,265 CTPA studies performed during the pre-intervention period, ordered by 25 unique providers.

1,582 studies were ordered during the final pre-intervention year, with 174 significant for positive PE findings, positivity rate 11.00%. In the post-intervention year, 1,339 CTPA studies were ordered for suspected PE, representing a 15.36% decrease compared to the final pre-intervention year. Of these, 183 were significant for positive PE findings, for an overall positivity rate of 13.67%. Thus, compared to the final pre-intervention year, post-intervention period positivity rate increased significantly by 2.67% (P = 0.028).

### Secondary Outcomes

Cumulatively during the baseline period, provider-specific positivity rate was normally distributed with a mean of 11.49%, standard deviation of 4.16%, and range from 4.59% to 20.00%. Mean provider positivity rates further stratified by year are shown in Figure 1.

**Figure 1.**
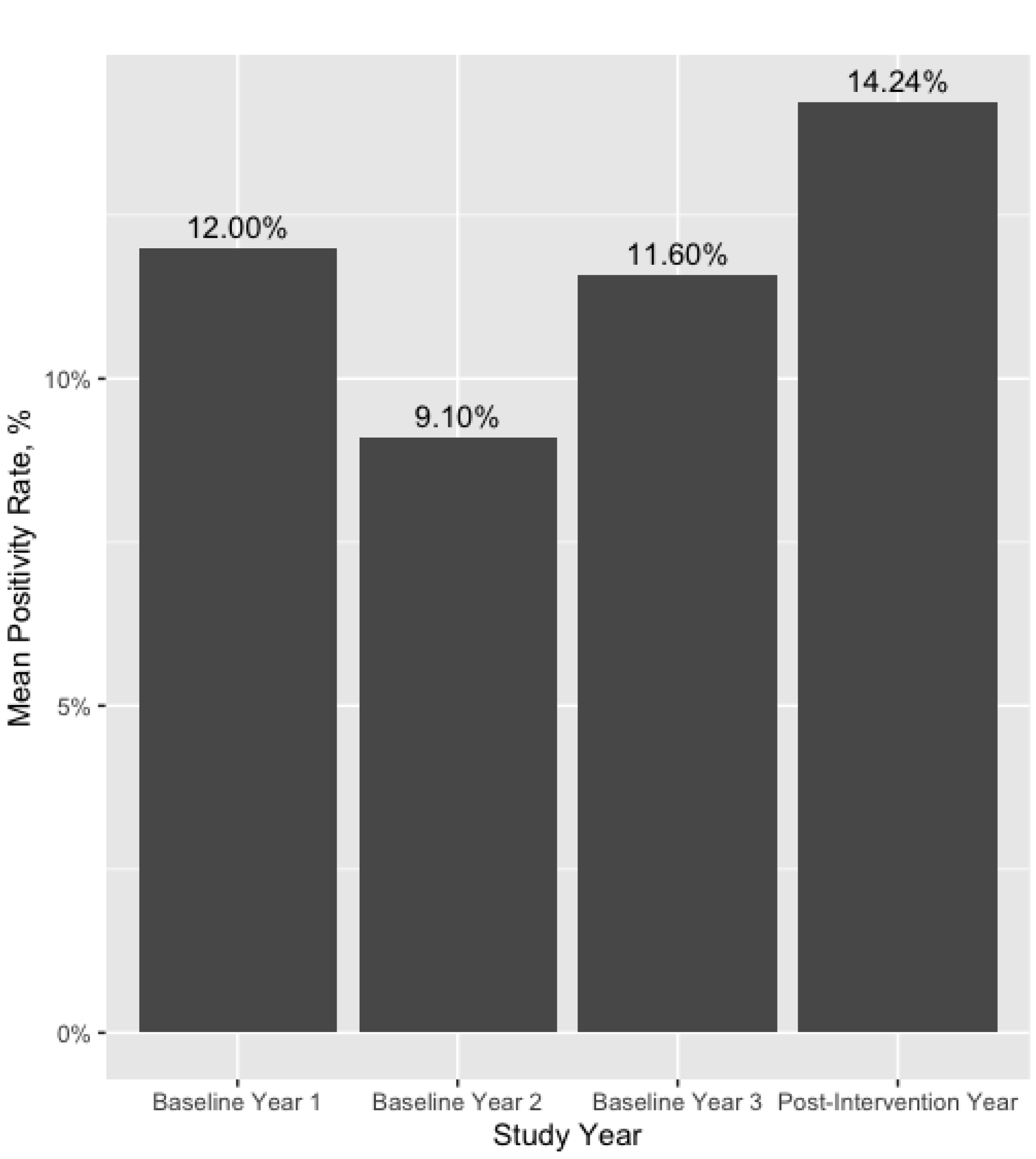
Mean provider positivity rate by study year

Additionally, provider-specific positivity rate in the post-intervention year was also normally distributed with mean of 14.24% and standard deviation of 7.32%. Range was 31.81% and, of note, one provider had a positivity rate of 0%. Figure 2 summarizes the differences in provider positivity rate between the pre- and post-intervention period. Table 1 further stratifies these results by individual provider. Compared to the preintervention period, mean provider positivity rate increased by 2.75%.

**Table 1.** Provider-specific Positivity Rate Before and After Intervention.

| Provider | Total Ordered | Positivity Rate, Baseline Period | Positivity Rate, Post-Intervention Period |
| --- | --- | --- | --- |
| A | 77 | 9.52 | 25.00 |
| B | 83 | 13.09 | 18.18 |
| C | 90 | 12.82 | 31.81 |
| D | 99 | 8.45 | 15.38 |
| E | 115 | 20.00 | 22.73 |
| F | 117 | 16.86 | 10.52 |
| G | 120 | 9.09 | 10.00 |
| H | 126 | 9.09 | 11.54 |
| I | 130 | 14.63 | 4.00 |
| J | 175 | 10.00 | 23.50 |
| K | 179 | 11.53 | 19.30 |
| L | 184 | 13.33 | 17.65 |
| M | 201 | 17.91 | 12.5 |
| N | 210 | 18.88 | 5.55 |
| O | 220 | 9.09 | 22.86 |
| P | 223 | 16.08 | 8.33 |
| Q | 243 | 8.8 | 0 |
| R | 250 | 8.20 | 8.00 |
| S | 251 | 6.70 | 14.81 |
| T | 281 | 9.09 | 16.28 |
| U | 308 | 11.56 | 19.40 |
| V | 379 | 15.35 | 9.09 |
| W | 456 | 4.59 | 6.94 |
| X | 497 | 5.81 | 12.16 |
| Y | 590 | 6.74 | 10.44 |
| **Mean** |  | 11.49 | 14.24 |
| **Standard Deviation** |  | 4.16 | 7.32 |

**Figure 2.**
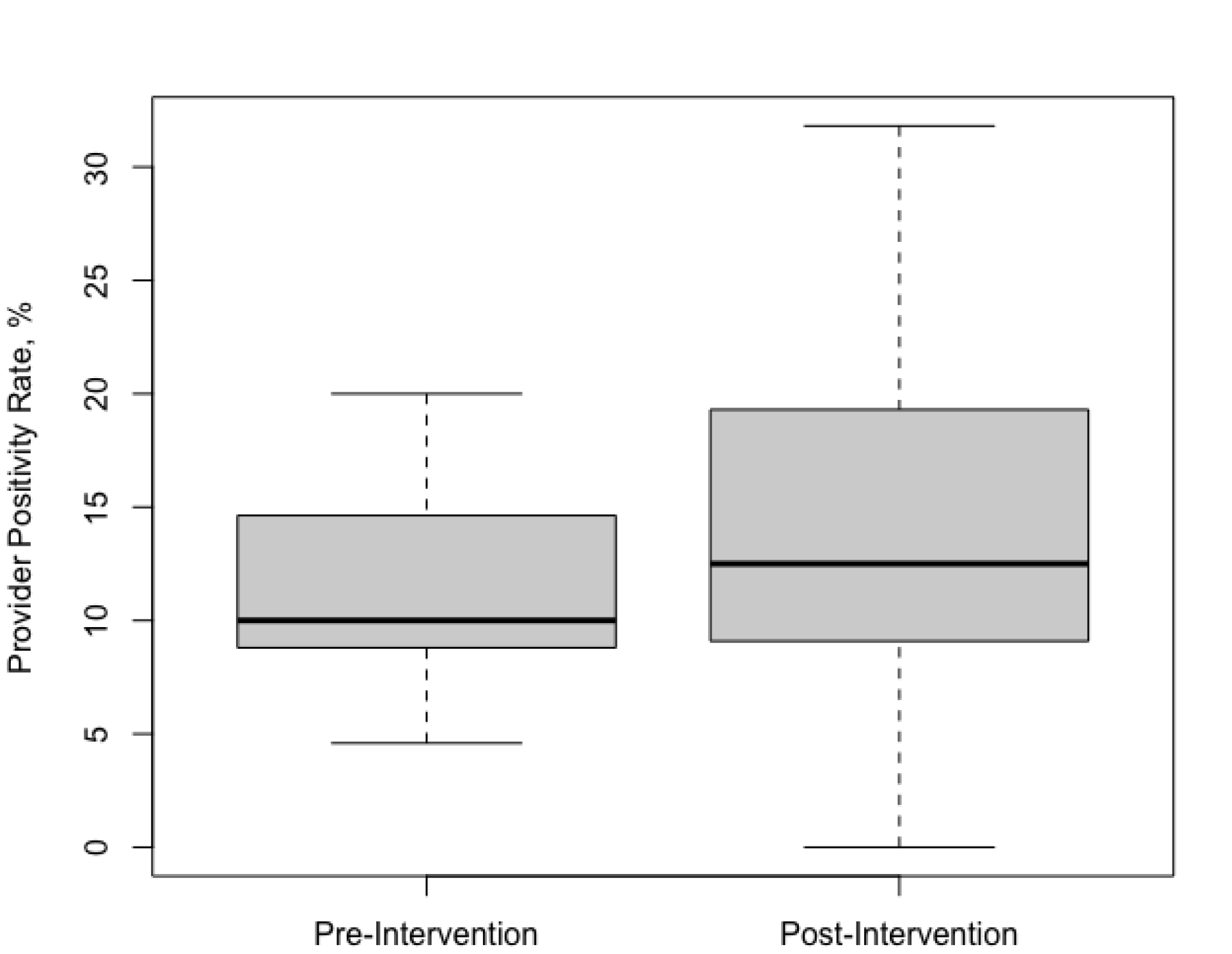
Provider Positivity Rate Before and After Intervention

Finally, as a post-hoc analysis, we identified providers who experienced high growth in positivity rate between pre- and post-intervention periods and compared baseline positivity rates. We defined high growth as >= 7.70%, the 75^th^ percentile in our study cohort. We then compared these providers to those who did not achieve high growth. 7 providers (28.00%) achieved high growth; 18 (72.00%) did not. Mean pre-intervention positivity rate was 10.17% with SD 2.02% in the high growth group compared to mean of 11.99% and SD of 4.80% in the non-high growth group. Notably, mean preintervention positivity rate did not significantly differ between groups.

## Discussion

CTPA utilization for evaluation of suspected PE is increasing across the United States. A substantial portion of ordered CTPAs are not justified by validated guidelines, and this increased ordering has led to a decrease in positivity rate, and thus reduced its value as a diagnostic tool^5^. In this study, we designed and implemented a novel, multimodal intervention combining provider education and individual performance analysis, which successfully increased CTPA positivity rate.

Our primary outcome was the significant 2.67% increase in positivity rate between the final baseline and post-intervention years, indicating that at the department-level, our intervention was effective. As noted, CTPA studies are costly and confer significant patient medical risk. At our institution, a 15.36% reduction in CTPA ordering during the post-intervention year represented cost savings exceeding $200,000 and a cumulative decrease in patient radiation exposure by up to 2,430 mSv. Furthermore, as CT scanner time is a scarce resource, the resultant reduction in CTPA ordering from our intervention also freed up scanners for other patient imaging needs.

Two important provider-specific results add nuance to our findings. First, In the preintervention cohort, provider positivity rate during the baseline period varied significantly, with a standard deviation of 4.16%. This finding is expected, as individual provider ordering practices can vary substantially. However, despite receiving an educational intervention aimed at better standardizing ordering practices, in the postintervention year, inter-clinician variability increased, with positivity rate standard deviation increasing to 7.31%. Second, in our post-hoc analysis, baseline year positivity rates did not significantly differ between providers who achieved high positivity rate growth, and providers who did not. In other words, providers with the greatest improvement in positivity rates did not demonstrate poorer adherence to guidelines at baseline.

An important strength of our study is that, as all providers practiced at the same emergency department, they were not subject to significant differences in institutional policy, patient population, or other external factors that could have influenced results. Differences in ordering practices, and consequently positivity rates, can therefore primarily be ascribed to physician-specific factors. Thus, our provider-specific findings emphasize two key concepts that be used to guide future interventions. First, educational and performance analysis interventions may have variable impact on providers. As such, tailoring interventions on a provider-specific basis may further increase guideline adherence and positivity. Second, individual improvement in guideline adherence and positivity rate is likely a function of physician-level factors other than baseline positivity rate.

Interventions aimed at improving provider guideline adherence and positivity rate can largely be stratified into three main categories: clinical decision support, education, and individual performance review.

Clinical Decision Support (CDS) systems have been well studied; however, results have varied substantially, with multiple noting nonsignificant changes to positivity rates^6-8^.

Notably, such CDS systems are often voluntary, and consequently may be ignored by ordering providers, reducing their value^9^. Mandatory CDS has shown promise, with Soo Hoo and colleagues achieving consistent, >=15% positivity rates for seven consecutive years since implementation^3^. Prior studies evaluating effectiveness of educational interventions have focused primarily on outcome metrics other than positivity rate; however, Booker and colleagues found a primarily educational intervention to have no significant increase on positivity rate^10,11^.

Individual performance analysis interventions remain understudied compared to the other two intervention types; however, neither Raja et. al nor Salehi et. al achieved significant increase in positivity rate through this intervention type alone^12,13^. However, one notable difference between these studies and ours, is that while our intervention provided only one-time feedback to providers, these studies provided periodic reports. This suggests that ordering providers may be more likely to adhere to guideline-related feedback during the first post-intervention year if received only once, potentially due to feedback fatigue.

While our intervention focused primarily on education and performance feedback, Soo Hoo and colleagues correctly point out that both those modalities can be cost and time-intensive endeavors^3^. As such, frequent use of those methods alone may constitute a substantial resource drain while yielding diminishing results. Ultimately, a multimodal, individualized approach combining judicious, education and performance analysis with effective clinical decision support may prove the highest-value intervention and certainly warrants further evaluation.

### Limitations

Our study was a single-center quality improvement project. As such, participants were not randomized; rather, all ordering providers received the intervention. From an experimental standpoint, a completely prospective, randomized controlled trial would be more robust and mitigate potential selection bias. Furthermore, the only formal education that providers received regarding PE criteria and CTPA use guidelines was our intervention. However, some ordering providers, may have received additional education on these topics through alternative means, such as self-directed learning, and consequently changed their ordering practices. Finally, our intervention did reduce the number of CTPAs ordered for suspected PE, and it is possible that the reduced number of orders led to missed or delayed PE diagnoses. However, the risk of missed diagnosis was well-considered during development of the clinical prediction rules. Thus, while improved guideline adherence by ordering providers may have conferred an increased risk of missed diagnosis during the post-intervention period, that risk was ultimately balanced by reduced inappropriate CT use.

## Conclusions

A multimodal intervention involving individualized performance analytics and education effectively increases diagnostic yield of CTPA for suspected PE in the emergency setting. Optimizing CTPA ordering reduces patient radiation exposure, provides substantial cost savings, frees scarce healthcare resources, and increases the value of the imaging tool.

## Data Availability

The authors declare that they had full access to all of the data in this study and the authors take complete responsibility for the integrity of the data and the accuracy of the data analysis. The clinical data used to support the findings of this study are restricted by the Baylor College of Medicine Institutional Review Board in order to protect patient privacy. Data are available from Eric M. Rohren for researchers who meet the criteria for access to confidential data.

